# Investigating pathways of vancomycin-resistant *Enterococcus* (VRE) contamination and transmission in intensive care units: a prospective genomic surveillance study

**DOI:** 10.64898/2026.07.28.26359040

**Authors:** Tierney O’Sullivan, Windy D. Tanner, William Brazelton, Karim Khader, Brian Orleans, Candace Haroldsen, Matthew H. Samore, Michael Rubin, Lindsay T. Keegan

## Abstract

**Background:** Vancomycin-resistant *Enterococcus* (VRE) species are common healthcare-associated pathogens that cause difficult-to-treat infections. Whole genome sequencing of patients has revealed a substantial burden of patient-to-patient VRE transmission in hospitals, with patients in intensive care units (ICUs) at particularly high risk of acquisition. However, few studies adequately characterize the pathways of VRE transmission between patients in acute care settings—a necessary step to identify current gaps in infection prevention practices. By harnessing genomic clustering analyses of whole genome sequences of VRE isolates from patients, environmental surfaces, and healthcare providers (HCP) in ICUs, we aim to reconstruct indirect pathways of pathogen movement to identify patterns of VRE spread and opportunities for transmission prevention.

**Methods and Findings:** We collected daily samples (N = 6848) from ICUs in two hospitals over 13 weeks from four main sampling sources: patients, HCP hands, patient rooms, and shared surfaces. Samples were cultured on selective media and sent for whole genome sequencing (WGS). We used genomic thresholds to identify clusters of related VRE isolates and distinguish unrelated isolates. VRE was detected in samples from 20 out of 322 unique occupant-stays (6.22%). VRE isolates were detected from all sampling sources except for shared surfaces. A total of 44 unique VRE isolates were identified, 43 *Enterococcus faecium* (VREfm) and one *Enterococcus faecalis* (VREf). Two distinct patterns of VREfm spread were observed: 1) an outbreak setting with observed patient-to-patient transmission and low VRE genomic heterogeneity, and 2) high VRE genomic heterogeneity and pathogen movement between occupant-stays facilitated by persistent HCP and environmental contamination, but no observed transmission events. VRE detection probabilities were not significantly different between occupant-stays in outbreak and non-outbreak settings (OR = 0.63, 95% CI (0.23, 1.83), *p* = 0.32). However, including VRE isolates from non-patient samples increased the number of occupant-stays with VRE detection from 6 to 20, a 3.3-fold increase, as compared to patient samples alone. Including non-patient samples also increased the number of VRE multi-isolate genomic clusters detected by 7-fold. Our findings are limited because sampling was primarily conducted in ICUs. Due to the combination of short ICU stay durations and imperfect test sensitivity, VRE transmission events may have been underdetected.

**Conclusions:** Our findings characterize the complex nature of VRE transmission pathways in ICU settings. Even without an ongoing outbreak, we found substantial evidence of VRE movement between occupant-stays, facilitated by a combination of HCP hands and environmental surfaces. This study highlights the importance of environmental sampling for understanding VRE transmission potential, which is likely to be underestimated using patient sampling alone. We recommend that future studies incorporate follow-up sampling after discharge to better understand the true burden of transmission.

## Introduction

Vancomycin-resistant Enterococcus (VRE) species, primarily *Enterococcus faecium* (VREfm) and *Enterococcus faecalis* (VREf), are major contributors to the burden of healthcare-associated infections (HAIs) worldwide.^1^ In the US alone, VRE infections cause an estimated 54,500 hospitalizations, 5,400 deaths, and over $500 million in attributable healthcare costs annually.^2^ Beyond their direct clinical and economic impacts, VRE species also serve as reservoirs of antibiotic-resistant plasmids, which facilitate the spread of antimicrobial resistance genes.^3^ VRE contamination is widespread in healthcare settings,^4^ driven in part by traits such as viability on environmental surfaces for days to months,^5^ and increasing tolerance to handwash alcohols^6^ and surface disinfectants.^7^ Reducing VRE transmission between patients is critical to limiting preventable adverse events and their associated healthcare costs, and perhaps more importantly, slowing the spread of antimicrobial resistant infections broadly.

Genomic tools are increasingly used to enhance our understanding of the prevalence, contamination, transmission, and importation of healthcare associated pathogens, including VRE, in hospital settings.^4,8–10^ In fact, recent findings from prospective and comprehensive genomic surveillance studies demonstrate that healthcare-associated transmission is higher than previously estimated by clinical isolates alone, with 60-80% of patients estimated to have acquired VRE during their hospital stays, rather than prior to admission.^9–11^ Intensive care units (ICUs) in particular are identified as hot spots of VRE transmission.^9^

While these recent studies highlight the ongoing problem of healthcare acquisition, and how current infection prevention and control practices struggle to prevent VRE spread, they are not designed to reveal how patient-to-patient transmission is occurring. In acute care settings with little to no direct patient-to-patient contact, characterizing the relative contributions of indirect transmission pathways is critical. Current studies of healthcare-associated pathogens suffer from one of two main limitations that prevent them from adequately characterizing these indirect transmission pathways. First, many studies assess the relatedness of VRE isolates with methods that lack sufficient discriminatory power to reconstruct indirect transmission pathways and to determine who-infected-whom^12^ (*e.g.*, pulsed-field gel electrophoresis or multilocus sequence typing [MLST]).^4,13–17^ Second, the studies that do incorporate more granular genomic methods based on whole genome sequencing (WGS) are either limited to patient isolates alone,^9,12,18,19^ or do not sample environmental surfaces or healthcare providers (HCP) sufficiently to holistically characterize the role of indirect pathways of transmission between patients.^10,20,21^

VRE species are ideal candidates for WGS methods to characterize the role of indirect transmission dynamics in healthcare settings for epidemiologic and genomic reasons. Epidemiologic reasons include previously mentioned characteristics such as its prevalence in patient populations, viability on environmental surfaces, and tolerance of hand sanitizers. Genomically, compared with other healthcare-associated pathogens with similar environmental persistence qualities, such as *Clostridioides difficile* (*C. difficile*), VRE species have among the fastest genome-wide evolutionary rates.^22^ This fast evolutionary clock aids in genomic epidemiology inference, with rapidly accumulating mutations providing more clarity between related local transmission chains and unrelated imported strains.^23^

To address the limitations of prior genomic epidemiology studies of healthcare-associated pathogens in characterizing indirect transmission pathways, we use a longitudinal study design of WGS from comprehensive patient, HCP hand, and environmental sampling from patient rooms and shared surfaces in ICUs in two hospitals. Results from the dense sampling collected in this same study for another pathogen, *C. difficile*, indicated that detection of cryptic transmission through HCP hands and environmental surfaces was 3.6 times higher than what would be estimated from patient sampling alone.^24^

The longitudinal sampling design allows us to distinguish between evidence of VRE transmission, acquisition, importation, and movement. Using genomic and epidemiologic data from a prospective longitudinal sampling design, we can reconstruct VRE transmission and movement between intensive care unit patients, HCP hands, and environmental surfaces to identify patterns of VRE spread and opportunities for control.

## Methods

This study was approved by the University of Utah Institutional Review Board. All study participants, patients and HCPs, provided consent prior to sample collection.

### Study Location and Timing

The study was conducted in 2018 over a period of 13 weeks in ICUs at two different facilities, denoted as hospitals A and B.^24^ Study sites included a 20-bed cardiovascular ICU (CV-ICU) in hospital A, which was sampled for 56 consecutive days, and a 10-bed ICU in hospital B, sampled for 35 consecutive days.

### Sampling Design

To characterize the indirect pathways of VRE transmission in intensive care settings, we collected 6848 samples from four broad categories of sample sources: patients, HCP hands, patient room surfaces, and shared environmental surfaces. Patients who were present at the time of sampling and consented were sampled daily from up to three specific body sites: a) axilla, 2) groin, and 3) either perianal or stool. All patients were eligible for sampling upon admission to the ICU and remained eligible up to one day following discharge from the ICU. If a patient was transferred to another unit but remained in the facility, they were followed for patient sampling for one additional day. HCP hands, either gloved or bare hands, were sampled upon exiting a patient’s room in the ICU and before hand hygiene.

For patients not on transmission-based contact precautions, patient room samples were taken as composites from each of three zones: 1) near-patient (e.g., bedrails), 2) HCP high-touch area (e.g., computer cart), and 3) toilet area. For patients on transmission-based precautions, individual patient room surfaces were sampled separately. Terminal cleaning was performed after each patient was discharged or transferred from a room before the next occupant was admitted. Shared surfaces on the ward were also sampled daily, including the nurses’ station and shared mobile equipment not affiliated with a unique occupant-stay. See Supplement Tables S-1 and S-2 for a complete description of the environmental sampling locations and their respective surface areas. See Keegan et al.^24^ for further details on the sampling methods.

The timing and location of each sample collection were recorded. For simplicity, all categories of transmission-based precautions (contact, droplet, etc.) are aggregated. All patients were given a unique identifier (e.g., Patient A004), and all samples associated with their occupant-stay were linked to that identifier, including samples from their body sites, samples from HCPs leaving their room, and samples from surfaces in the room they occupied at the time of sampling. If a patient was discharged from the facility and then readmitted, they were given a new patient identifier. When patients were transferred between rooms within an ICU, the assigned occupant-stay identifier followed the patient, resulting in some occupant-stays involving more than one patient room.

### Microbiology Methods

Environmental surfaces and HCP hands were sampled with 3M cellulose sponges (3M^TM^ Sponge-Stick with neutralizing buffer, 3M, Sparks, MD). Patient body sites were sampled using Regular Flocked BD Liquid Amies Elution Swabs (Eswabs) (Becton Dickinson and Company, Sparks MD). To target VRE, all samples were cultured on selective ChromID agar supplemented with vancomycin or in Bile Esculin Broth with a vancomycin disk.

To identify morphologically-distinct enterococci colonies to species-level, colonies were streaked to isolation and identified with MALDI-TOF. Isolates were frozen and later grown up in pure culture to test for vancomycin susceptibility by disk diffusion (30 µg vancomycin disk). See Supplemental methods for detailed microbiologic methods.

### Genomic and Clustering Analysis

All morphologically distinct suspected *Enterococcus* colonies were isolated, sent to the Huntsman Cancer Institute High-Throughput Sequencing laboratory, and sequenced on an Illumina Nova-Seq 6000 platform at approximately 50x coverage. All VREfm WGS isolates were identified to clades using fastANI (version 1.34)^25^ against clade-specific reference genomes (Table S-4). To classify each isolate by MLST, we queried genome assemblies against the PubMLST *Enterococcus faecium* and *Enterococcus faecalis* databases.^26^ All VRE species with more than one isolate, as identified by MALDI-TOF, were eligible for genomic clustering analysis.

Based on recent research identifying optimized genomic approaches for identifying probable VRE transmission,^12^ we used split kmer analysis (SKA, version 1) ^27^ as the primary method to establish clusters of the VRE isolates. Raw reads were quality-trimmed (cutoff = 30) and pairwise distances were calculated between isolates, with parameters for kmer length set to 19 and minor allele frequency set to 0.01.^28^ Clustering links were established between each pair of isolates that matched two *a priori* criteria: a) Jaccard Index ≥ 0.9, which measures the proportion of quality-trimmed kmers that matched between isolates, and b) the number of single nucleotide variants (SNVs) between isolates ≤ 7.^12^ Genome clusters were constructed using a single-linkage clustering algorithm. We distinguish between distinct VRE clusters based on our genomic *a priori* thresholds; some isolates formed multi-isolate clusters, whereas others did not and are referred to as singletons.

To assess the stability of the clusters that were formed using SKA with the above *a priori* thresholds, we conducted multiple sensitivity analyses. We first assessed the sensitivity of the genomic distance calculation method on cluster stability. We compared clusters formed with the primary method (SKA) with clusters formed using an alignment-based method against a reference genome to calculate pairwise SNV distances (Table S-4). We then assessed the robustness of the SKA results from our clustering analysis by testing a set of SKA algorithm parameters and alternative SNV thresholds found in the literature for VRE transmission in healthcare settings.^24^ For detailed information about the alignment-based methods and the range of methods tested in the sensitivity analyses, see the Supplemental Methods.

### Statistical Methods

We used descriptive statistics to characterize VRE detection rates from the different sampling environments and facilities, as well as the timing to first VRE detection. To test for differences in the distributions of the length of occupant-stays with VRE isolates vs. those without VRE isolates, we used a Wilcoxon rank sum test. To test for the difference in VRE detection between facilities, sampling environment types, and in glove vs. bare hand HCP hand samples, we used a Fisher’s exact test and report conditional odds ratios (OR) and 95% confidence intervals (CI). P values less than 0.05 are deemed significant. All statistical analyses were conducted using R version 4.6.0.^25^

### Terminology

We used epidemiologic and genomic data as evidence to distinguish between acquisition, importation, transmission, and pathogen movement. For patient samples, we broadly define VRE acquisition, consistent with other studies,^29,30^ as patients with two or more days of samples without VRE detection followed by VRE detection. This definition is solely based on timing of detection, regardless of whether the patient’s VRE isolate genomically clusters with isolates already detected on the ward. VRE importation was defined as any patient with VRE detection within their first two days on the ward. We define VRE transmission as a special case of VRE acquisition. Transmission is defined as patient acquisition of VRE that genomically matches previously detected VRE isolates on the ward, regardless of whether it was previously detected in a patient or non-patient sample. Because we are interested in the pathways of potential indirect transmission, we include both patient and non-patient samples to investigate routes of pathogen movement in the ICUs. We define pathogen movement as isolates from the same genomic cluster detected across multiple sampling locations, either different environmental surfaces, patients, or HCPs.

## Results

### VRE detection

There were 322 unique occupant-stays across the two ICUs during the study period: 225 from hospital A, and 97 from hospital B. Of the 322 unique occupant-stays, 6.6% had VRE isolates from any sample environment affiliated with their occupant-stay (6.6%, 20/322 total; 5.3%, 12/225 in hospital A; 8.2%, 8/97 in hospital B). While the percent of occupant-stays with VRE isolates was lower in hospital A, the difference was not statistically significant (OR = 0.63, 95% CI = (0.23, 1.83), *p* = 0.32).

Each specific sampling environment was eligible for daily sample collection, but actual sampling coverage varied across hospitals and environments. Sampling coverage was lowest for patient body site sampling, which required patients to be present at the time of sampling and to consent. 56.2% of patients received at least one body site sample during their occupant-stay, though patients in hospital B had higher coverage (56.2% 181/322 overall; 51.1%, 115/225 in hospital A; 68.0%, 66/97 in hospital B). Sampling coverage of patient rooms was higher, with 86% of the 322 unique occupant-stays with at least one room environment sample taken (86.0%, 277/322 overall; 84.9%, 191/225 in hospital A, 88.7% 86/97 in hospital B). The proportion of patients with HCP hand samples affiliated with their occupant-stays was also high, with 94.4% coverage (304/322 overall; 95.6%, 215/225 in hospital A; 91.8%, 89/97 in hospital B).

Of the 6848 samples collected, 41 samples yielded a total of 44 unique VRE genomic isolates (0.60%; 22 samples yielded 25 unique isolates from hospital A; 19 samples, 19 unique isolates from hospital B). Because all morphologically distinct colonies from each sample were sent for WGS, three samples yielded two distinct VRE isolates. Of the 44 total isolates, 43 were *E. faecium,* and 1 was *E. faecalis* (recovered from hospital B).

The highest overall VRE detection rate across the four main sampling environments was the patient body site samples, with 0.9% of patient body samples yielding VRE (overall: 11/1291; hospital A: 0.2%, 2/896; hospital B: 2.3%, 9/395). This was followed by the patient room environment, with 0.7% (19/2654) VRE detection, and HCP hand samples at 0.5% (11/2402). None of the shared environment samples collected from surfaces unaffiliated with occupant-stays (e.g., shared equipment, nurses’ station) yielded any VRE isolates (0/501, 0%). Using patient body samples as the reference, differences in VRE detection rates for HCP hands (OR = 0.55, 95% CI = [0.23-1.35], *p* = 0.20) and patient room surfaces (OR = 01.09, 95% CI = [0.53-2.36], *p* = 0.87) were not significant. However, sampling effort and VRE detection rates varied across sample location types and healthcare facilities (Figure S-1, Table S-6).

Of the 181 patients with body site samples collected during their occupant-stay, 6 had VRE detection from at least one patient body site (3.3%, 6/181 total; 1.7%, 2/115 in hospital A; 6.1%, 4/66 in hospital B). Disaggregating the patient body site collections into more granular sample locations, VRE was detected in groin and perianal/stool samples but not in axilla samples. Perianal/stool samples from hospital B yielded the highest rate of VRE detection of any sample location at either facility, at 6.4% (7/109 samples), compared to hospital A perianal/stool samples at 0.9% (2/221 samples). Only two out of 435 (0.5%) groin samples yielded VRE, both of which were from hospital B. For the two patients with VRE detected in groin samples, on the collection days of the positive groin samples, both patients also had VRE detected in their perianal/stool samples.

Of the 277 occupant-stays with patient room samples collected, 10 had VRE detection during their stay (overall: 3.6%, 10/277; A: 3.1%, 6/185; B: 4.6%, 4/86). Of the 19 patient room samples that yielded VRE, 12 were from hospital A and 7 from hospital B. When we disaggregated patient room surfaces by facility and specific surface type, we found distinct patterns of VRE detection across the two facilities. The highest rate of VRE detection was in the HCP touch area (Zone 2) samples in hospital A with 1.7% positivity (4/236), whereas zero samples yielded VRE in the HCP touch area (Zone 2) in hospital B. Hospital B’s highest zones for environment VRE detection rate were the near patient area (Zone 1) and the patient toilet area (Zone 3), which were tied at 1.1%. Interestingly, none of the environmental samples from rooms of patients on contact precautions yielded VRE isolates. Because of the difference in room layouts, patient room sinks were tested more often in hospital B than in hospital A, yielding a 0.5% (1/184) VRE detection rate in hospital B, and no VRE detections in hospital A (0/12).

Of the 304 patients with HCP hand samples affiliated with their occupant-stays, 9 had VRE detection (2.9% overall; 2.8%, 6/215 in hospital A; 3.4%, 3/89 in hospital B). Of the HCP hand samples, VRE detection rates were similar across facilities, with hospital A having roughly 3 times the number of HCP hand samples collected (A: 8/1738, 0.5%; B: 3/664, 0.5%). There were also similar VRE detection rates among HCP hand samples from gloved (overall: 6/1221, 0.5%; A: 5/953, 0.5%; B: 1/268, 0.4%) or bare hands (overall: 5/1160, 0.4%; A: 3/772, 0.4%; B: 2/388, 0.5%) in both facilities and were not statistically different overall (*p* = 1).

The proportion of patients on any type of transmission-based precautions varied over time, with overall higher rates in hospital A (mean daily proportion of patients on precautions = 9.5% in hospital A; 3.2% in hospital B; Figure S-2). None of the 20 patients with VRE isolated from samples from their occupant-stay were on contact precautions, including the six patients with VRE detected from body-site sampling.

The average ICU length of stay for patients in the study was 3.8 days (median = 2, IQR = 1-4 days) overall. Patients in hospital A had a longer length of stay (mean = 4.2, median = 2, IQR = 1-4 days) than in hospital B’s ICU (mean = 2.6, median = 1, IQR = 1-2 days). Patients with VRE isolates affiliated with their occupant-stay had a longer length of stay (mean = 7.5, median = 4, IQR = 1.75-9.25 days) than those without VRE detections (mean = 3.5, median = 2, IQR = 1-4 days). Results of a Wilcoxon rank-sum test indicate that the stay lengths for patients with and without VRE detection differ significantly (*p* = 0.009).

For the 20 occupant-stays with VRE detection, the average time between ICU admission and the first VRE detection was 3.7 days (median = 1, IQR = 1-3.25 days). The first sampling environment to yield VRE was from patient samples in 25% of occupant-stays (5/20), room samples in 35% (7/20), HCP hands in 30% (6/20), and a tie between HCP hands and patient room in the remaining 10% (2/20) (Figure S-3). For occupant-stays with the first VRE detection from patient body sites, the mean time to detection was 1.8 days, whereas if the first VRE detection was from either HCP hands, the patient room, or both, the mean time to detection was 4.3 days.

### Genomic classification

All VREfm isolates were classified in Clade A1, an *E. faecium* clade known for adaptations to healthcare settings. MLST using 7 loci identified 38 of the 43 *E. faecium* isolates in a total of 4 sequence types: ST18 (18.9%), ST412 (29.7%), ST664 (2.7%), ST967 (48.6%), or did not match a specific MLST (16.2%) (Figure 1, Figure 2, Figure S-4). Based on MLST classification, the VREfm isolates from hospital A represented a broader range of sequence types, including all four detected sequence types and isolates that did not match a known type. Hospital B only included isolates from ST18 and ST967, both of which were also detected in hospital A. Of the VREfm isolates that matched with a known sequence type (37/43, 86.0%), all belonged to the hospital-adapted clonal complex CC17. The six VREfm isolates from hospital A that did not match a known type all shared identical allelic profiles. The sole VREf isolate was classified as sequence type 36, which is typically isolated from animal sources rather than human clinical sampling.^22^

**Figure 1.**
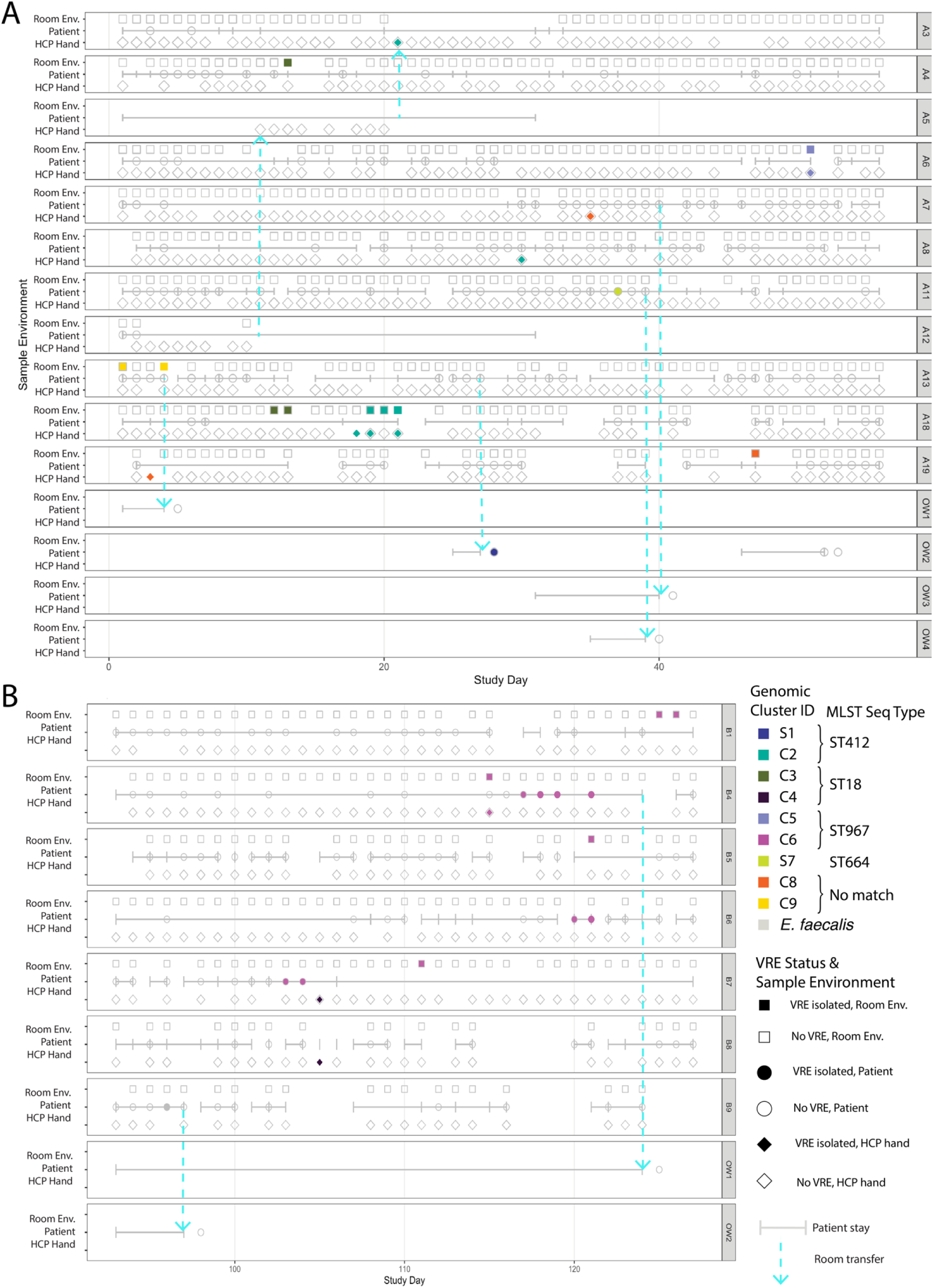
Study timeline from Hospital A and Hospital B. Each subplot represents a unique patient room that was used by patient with at least one VRE isolate affiliated with their occupant-stay, colored by VRE cluster ID, with shapes and subplot rows representing sample location. Sequential Room IDs indicate rooms that are adjacent to one another. OW = off ward, i.e. the patient was sampled outside of the ICU. Patient-stays are indicated with horizontal gray bars, and room transfers are indicated by dotted cyan arrows. For the genomic cluster IDs: S = singleton isolate, C = multi-isolate cluster

**Figure 2.**
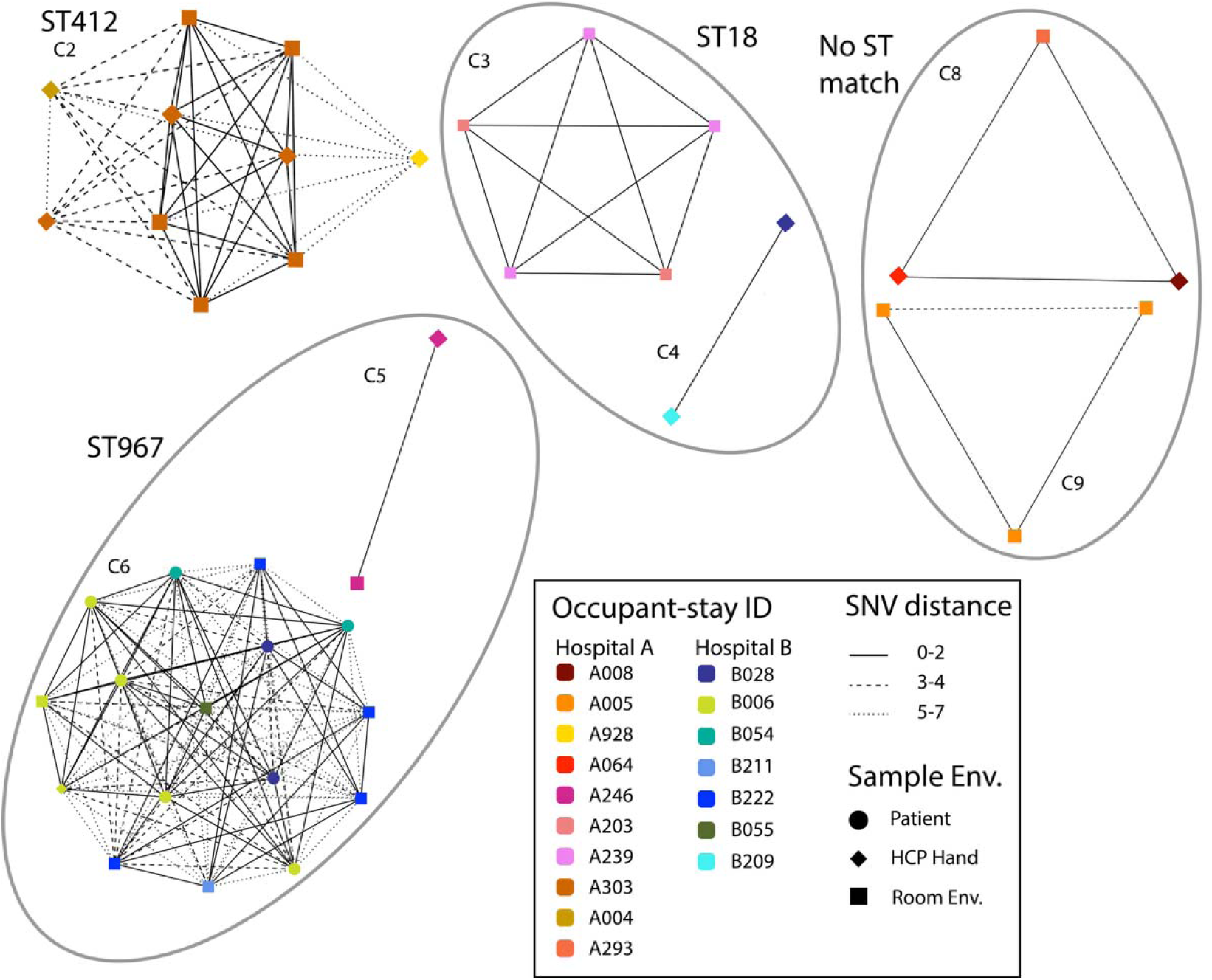
Network plots of VREfm (vancomycin-resistant Enterococcus faecium) multi-isolate clusters. Nodes represent individual VREfm isolates, which are colored based on the occupant-stay that the sample was affiliated with, and with shapes based on the sample environment. Edges that connect each node are weighted by the pairwise single nucleotide variant (SNV) distances. Singleton isolates that did not form clustering links are not depicted. Clusters are labeled by their matching multi-locus sequence type. Sequence types with more than one cluster are circled in gray.

All 43 VREfm genomic isolates contained the vancomycin resistance-conferring *vanA* gene, which was present on the plasmid assemblies. The *vanA* gene was not detected in the single VREf isolate, but it did include genes associated with virulence factors that were not detected in the VREfm isolates such as gelatinase (*gelE)*, and hyaluronidase (*hylB)*. The VREf isolate did have antimicrobial resistance genes, such as *vanZ*. The collagen adhesion protein virulence factor gene (*acm*) was present in all VRE isolates, regardless of species. The vancomycin resistance gene *vanB* was not present in any of the isolates.

### Genomic clustering

From the MLST, we found that VRE isolates from hospital B were more similar than those from hospital A; we refined this estimate using WGS clustering. Using the pairwise genomic linkage thresholds of 7 SNVs and 90% kmer matching, we reconstructed likely pathogen movement and transmission clusters. We found that all pairs of isolates from body sites of a single patient are genomically linked. However, not all samples associated with an occupant-stay (e.g., patient room surfaces, HCP hands) were genomically linked (Figure 3). The maximum genomic distance (number of pairwise SNVs) between samples from body sites of a single patient was 4 SNVs (Figure S-5).

**Figure 3.**
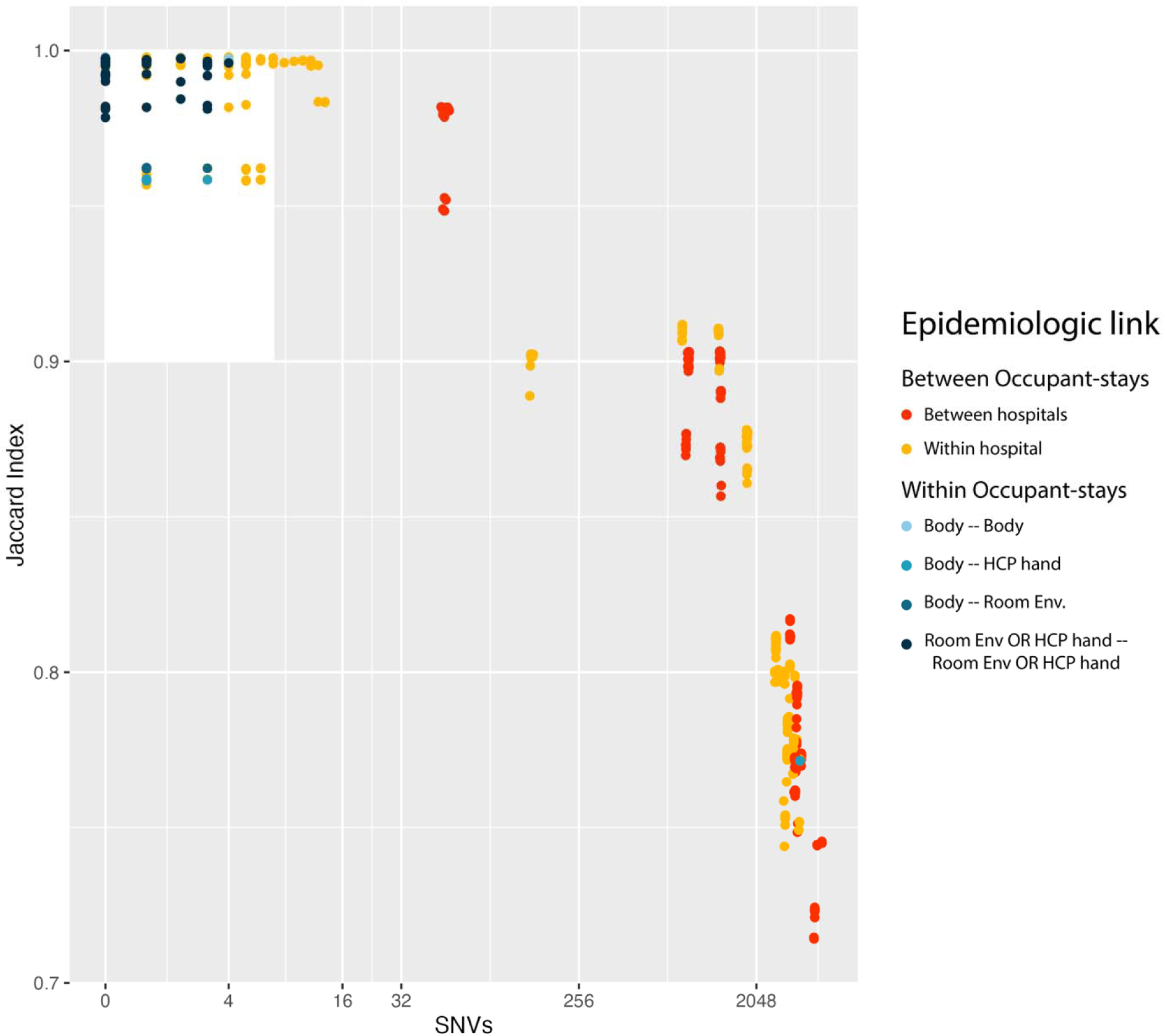
Scatterplot of pairwise genomic distances between all vancomycin-resistant Enterococcus faecium (VREfm) isolates calculated using split kmer analysis (SKA). Pairwise genomic distances are calculated using two metrics: single nucleotide variants (SNVs) (depicted along the x-axis, on a log scale), and the Jaccard index, or the proportion of split kmers that matched between isolates (y-axis). The white rectangle in the upper left corner indicates the pairs of isolates that formed clustering links according to the a priori cutoffs (7 SNVs, 0.90 Jaccard Index). All points represent a pair of isolates, and are colored by the epidemiologic link between them, with pairs of isolates from samples affiliated with different occupant-stays in the warm color scale and pairs of isolates taken from within a single occupant-stay depicted in the blue color scale.

From the 43 *E. faecium* isolates, we identified nine distinct genomic groups from our clustering analysis: seven distinct VRE multi-isolate clusters and two distinct singletons that did not form clustering links (Figure 1). All isolates in a given cluster were also identified as the same MLST sequence type. However, some sequence types contained isolates across multiple clusters, including isolates from both facilities. Isolates from patient body sites were only detected in three of the nine distinct genomic groups: one multi-isolate cluster and two genomically distinct singletons. Thus, including isolates from patient rooms and HCP hand samples increased the detection of unique VRE genomic groups by 3-fold, compared to restricting to patient samples alone. When restricting to the seven multi-isolate clusters (Figure 2), only one cluster involved isolates taken from patients (C6, hospital B). Thus, including non-patient isolates increased the detection of unique VRE genomic clusters by 7-fold.

Hospital A had the most genomically heterogeneous isolates, with seven unique VRE genomic groups from 25 isolates. Of these, three formed clusters that included isolates from multiple occupant-stays. This includes one patient who did not have a VRE isolate during their stay in the ICU but had a VRE isolate recovered from a body-site sample the day after transferring to a non-ICU ward within the same facility. Hospital A had the longest cluster detection duration, with intermittent detection of cluster C8 over 44 days (Figure 1). No pairs of samples taken from different hospitals formed genomic links (Figure 3).

Hospital B had less genomic heterogeneity in its VRE isolates. Separate from the sole *E. faecalis* isolate found in hospital B, genomic clustering identified only two distinct clusters from the 19 *E. faecium* isolates and no singletons. Both clusters involved isolates from multiple occupant-stays. Further, these two clusters involved isolates collected from adjacent rooms, a pattern not observed in hospital A.

Our clustering analysis indicates that terminal cleaning procedures were generally effective. Of the 10 occupant-stays with VRE detection on patient room surfaces, only one had room samples yield subsequent VRE isolates following their discharge or transfer (Room A18), though the subsequent VRE detection was unrelated genomically (clusters C2 vs. C3, Figure 1). This genomic evidence underscores the fact that the recurring VRE contamination of room surfaces in Room A18 between consecutive occupant-stays was not from ineffective terminal cleaning. However, there was some evidence of recurring cluster detection in the same room over multiple patients, including among consecutive (C6 in Room B7), and non-consecutive occupant stays (C8 in Room A19, Figure 1). Additionally, environmental samples from room A13 yielded VRE isolates from cluster C9, even though the patient occupying that room (Patient A005) consistently had no VRE detected in their body site samples or HCP hands. This indicates potential room contamination from the previous occupant, who was not sampled because their stay occurred prior to the study start date.

With the longitudinal study design and samples from patients and environmental surfaces, we can reconstruct likely transmission pathways and pathogen movement. In hospital A, we found evidence of VRE acquisition, but not transmission, as both patients acquired VRE that involved singletons not detected elsewhere. Their isolates were the only two singletons in the study, both from perianal samples (patients A054 and A073). We do, however, find evidence of pathogen movement in hospital A, with three VRE clusters (C2, C3, and C8; Figure 2) detected in samples across multiple occupant-stays. All three clusters detected across multiple occupant-stays in hospital A include VRE isolates from patient room samples, and two of the three include isolates from HCP hand samples (Figure 2).

In hospital B, we found evidence of both pathogen movement and transmission. The majority of the VREfm isolates (16/18) belonged to a single cluster involving samples from six unique occupant-stays (C6, Figure 2). The remaining two VREfm isolates also formed a cluster detected across multiple occupant-stays (C4). Cluster C4 involved two HCP hand samples from two unique HCPs in different occupant rooms on the same study day (both bare hands). Both of hospital B’s clusters included samples from HCP hands, while only the larger cluster (C6) involved patient samples or patient room samples. We found strong evidence of transmission between patients B028 and B006. Patient B028 was an imported case who had VRE isolated from a perianal sample collected on the day after admission. Patient B006 was repeatedly tested without VRE detection until day 23 of their stay, when their HCP hands and patient room samples yielded VRE isolates matching a cluster already detected on the ward. Patient B006 remained negative until two days later, when VRE was isolated from body site samples for the next four consecutive days of sampling. The other patient with VRE isolated from body site sampling was patient B054, who was readmitted to the ICU on study day 120 after their discharge approximately two weeks prior (Room B6, Figure 1). We never recovered any VRE isolates from any samples during their initial stay (not specifically identified here due to patient privacy), which lasted approximately one week and overlapped with Patient B028. We recovered cluster C6 VRE isolates from body-site samples from Patient B054 upon readmission, suggesting that they acquired VRE during their previous stay, but were discharged before testing yielded isolates.

### Sensitivity analysis

Our findings were robust to the genomic method used. Holding the SNV clustering threshold constant, the resulting clusters were identical across the genomic methods. However, relaxing the SNV threshold for establishing pairwise clustering links from 7 to 20 SNVs led to slightly different results in hospital A (Figure S-6). The 20 SNV threshold removed the distinction between two clusters (C8 and C9), establishing a new six-isolate cluster involving four unique occupant-stays. It does not involve any patient samples; only HCP hands and patient rooms are involved. It extends the longest cluster detection from 44 days to 46 days. All other clustering results remained unchanged.

## Discussion

By using dense longitudinal sampling from patients, HCPs, and a range of environmental surfaces in two ICUs, we observed two distinct patterns of VRE detection. The first pattern was characterized by multiple unrelated VRE clusters primarily detected on surfaces and HCP hands with limited patient colonization and no evidence of transmission. The second pattern involved a largely monoclonal outbreak with evidence of patient-to-patient transmission, likely mediated by the contamination of HCP hands and the patient room environment. In settings with surveillance-based patient sampling, the second pattern of patient-patient VRE transmission of a monoclonal outbreak would likely have been detected. However, the first pattern would not have been detected by traditional surveillance. With our culture-based methods, some patients had transient VRE detection: two of the four patients with VRE detected from perianal or stool samples had at least one follow-up perianal/stool sample yielding no VRE. Thus, even with surveillance-based patient testing, without repeated sampling, true VRE prevalence is likely to be underestimated. With either pattern, the true VRE prevalence and genomic heterogeneity, both in colonized or infected patients and on environmental surfaces, would be severely underestimated if relying on clinical samples alone. In both hospitals at the time of the study, clinical VRE detection required the patient to be placed on contact precautions; none of the patients with VRE isolates from their occupant-stays were on precautions, indicating that none of the patients from whom we recovered VRE were clinically detected during the study period.

We found that including HCP hand and environmental sampling led to a 3.3-fold increase in VRE detection in unique occupant-stays (6 patients with body samples, vs. 20 with any samples affiliated with their occupant-stay), similar to findings for *C. difficile* from the same study.^24^ The overall detection of VRE among patients was lower than most reported prevalence rates from surveillance studies,^31^ indicating that our observed importation, transmission, and contamination rates may also be lower than in typical settings.

While this study was not systematically designed to assess the effectiveness of transmission-based contact precautions, such as gowning and gloving, in preventing the spread of VRE, it did provide observational evidence suggesting that contact precautions may be protective against acquisition. None of the patients on contact precautions in either facility had any VRE isolates associated with their occupant-stays. It also provides evidence that patients without clinical VRE detection and therefore not on contact precautions may be acting as sources of VRE transmission. This cryptic transmission may be obfuscating the true effectiveness of contact precautions on VRE acquisition rates.^26^

The dominance of *E. faecium* isolated from our study is likely explained by the microbiologic culture methods, which selected for vancomycin-resistant strains. Rates *of E. faecalis* vs. *E. faecium* prevalence in healthcare settings vary by geographic region,^32^ but *E. faecium* is more likely to be vancomycin-resistant.^33,34^ Interestingly, the *E. faecalis* isolate grew successfully on selective media for vancomycin resistance, though it lacked the vanA and vanB genes.

Though the comprehensive sampling design was a strength of the study, it imposed some limitations due to feasibility constraints. One was the study duration (13 weeks total, 8 weeks in hospital A, 5 weeks in hospital B), which made it difficult to assess the true duration of clusters on the ward or long-term transmission dynamics, such as the length of transmission chains or final outbreak size. Additionally, sampling was mostly confined to patients, HCP, and environments within the ICU ward, and follow-up patient testing was conducted only one day after transfer for patients transferred to other non-ICU units within the facility. Patients were not followed up after discharge from the facility, and with most ICU length of stays less than 4 days, it is likely that patients who acquired VRE in the ICU were discharged before VRE would be detected. This is especially true for VRE, which has an estimated time between exposure and detection of 9 days.^35^ It is likely then that the observed number of VRE acquisitions is an underestimate.

Problems with VRE detection due to short lengths of stay are compounded by VRE test sensitivity. Sensitivity of VRE detection for culture-based genomic isolates ranges between 60 and 80%, depending on the method of collection (with sponge sticks having higher sensitivity than flocked swabs).^36^ We used multiple collection methods, so our testing sensitivities varied by sample environment. This may also explain the intermittent recovery of VRE from patient samples.

While clustering genomic isolates to establish putative transmission links based on an *a priori* genomic threshold may be sensitive to the genomic method and threshold choice,^28^ results from our sensitivity analysis indicate that our findings are robust to genomic methods, with perfect cluster replication across methods (i.e., kmer versus alignment-based). When we relaxed the SNV threshold from 7 to 20, we found that two clusters, each including three isolates, combined into a larger six-isolate cluster involving four unique occupant-stays. While this provided additional evidence of pathogen movement, neither of the two clusters that combined involved patient body isolates. Thus, our findings regarding patient acquisition were robust to the choice of SNV threshold. Although clustering results were generally robust, all methods we tested relied on substitution-driven measures of genomic distance and did not explicitly account for other mutational events (e.g., insertions, deletions, or structural variation). Future studies could explore more comprehensive genomic distance metrics that capture a broader spectrum of genomic change.

Our results provide suggestions for future studies looking at the indirect pathways of VRE transmission. We confirmed findings from other studies that MLST is too crude a metric for identifying putative transmission and pathogen movement.^12,28^ We confirmed that kmer-based genomic methods are sufficiently discriminatory for reconstructing putative transmission clusters, and we recommend these over alignment-based methods for efficiency.^12,28^ Stool or perianal samples had the highest sensitivity, and always yielded VRE when groin samples were positive, within the same cluster. Because we only had either perianal or stool samples from each patient, we are unable to compare detection rates between the two. However, stool sampling may be optimal for increasing coverage, as patients were more likely to consent to stool sampling.

Our findings raise questions about the standard definition of acquisition, which requires the first positive test to occur more than 2 days after admission, following negative admission testing. Based on this definition, we identified three acquisitions, but two did not match any other VRE genomic cluster on the ward. This indicates that 48 hours may be too strict a cutoff for differentiating importation versus acquisition. We acknowledge that the two observed acquisitions involving single isolates that did not cluster could be explained by incomplete sampling of all possible reservoirs, such as unsampled patients, night shift HCP, visitors, or other wards. However, with the time between exposure and VRE detection estimated at 9 days,^35^ we suspect these may have been importations instead. These findings provide evidence that a simple cutoff based solely on time may not effectively distinguish between acquisition and importation.

In conclusion, our study combines genomic surveillance with dense longitudinal sampling of patients, HCPs, and environmental surfaces to characterize the role of indirect pathways of VRE transmissions in intensive care settings. By including samples from indirect pathways, we revealed over three times as much pathogen movement between patient-stays as would have been detected from patient sampling alone. Even in settings without VRE outbreaks, VRE contamination of environmental surfaces and HCPs is common, with long-term detection outlasting single patient stay durations. Our findings point to complex intermittent positivity of multiple surfaces and HCP hands, rather than a single reservoir seeding transmission and movement. Our study was limited by focusing primarily on ICU testing and short lengths of stay. Future studies should expand the sampling after ICU discharge to better estimate how many transmission events are missed by within-facility testing.

## Data Availability

All de-identified data produced in the present study are available upon reasonable request to the authors

## Acknowledgements

We thank Rachel B. Slayton and L. Clifford McDonald for their contributions to the design of the parent study from which these data were derived. We also thank Rachel B. Slayton, L. Clifford McDonald, and Damon J.A. Toth for their insightful comments and constructive feedback on presentations of this research.

## Supplementary Material

### Section 1: Environmental Sampling Methods

**Table S-1.** Hospital A total surface areas of environmental surfaces and surface composites.

| Surface | Description | Total surface area of composite |
|---|---|---|
| Patient room Zone 1 | Composite of near-patient touch area surfaces – bedrails, over bed table, call button/remote; patient/provider touch overlap areas | 194 sq in |
| Patient room Zone 2 | Composite of healthcare personnel touch area surfaces – computer keyboard, scanner handle, room supply cart handles, IV pole high touch area | 300.5 sq in |
| Patient room Zone 3 | Composite of patient toileting area surfaces – toilet grab bars, flush handle, rinse spout. Alternatively: commode handles if commode present | 76 sq in |
| Patient over bed table | High touch area of patient over bed table (relevant precautions rooms only) | 132 sq in |
| Patient bedrails | High touch area of both patient bedrails (relevant precautions rooms only) | 108 sq in |
| Door touch areas | High touch areas of door – handles, door grab areas (relevant precautions rooms only) | 130 sq in |
| Sinks | Sink area – sink top, handles, soap dispenser button, hand sanitizer dispenser button | 114 sq in |
| Shared unit equipment | Surfaces of reusable shared unit equipment used in patient rooms | Variable |
| Shared nursing area | Composite of high touch areas in nurses station – two computer keyboards and two telephone handles | 140 sq in |

**Table S-2.** Hospital B total surface areas of environmental surfaces and surface composites.

| **Surface** | **Description** | **Total surface area of composite** |
|---|---|---|
| Patient room Zone 1 | Composite of near-patient touch area surfaces – bedrails, over bed table, call button | 173 sq in |
| Patient room Zone 2 | Composite of healthcare personnel touch area surfaces – computer keyboard, scanner handle, in-room medication dispenser, IV pole high touch area | 181 sq in |
| Patient room Zone 3 | Composite of patient toileting area surfaces – toilet grab bars, flush handle. More commonly VA patients used commodes. Commode handles sampled if commode present | 61 sq in |
| Patient over bed table | High touch area of patient over bed table (relevant precautions rooms only) | 124 sq in |
| Patient bedrails | High touch area of both patient bedrails (relevant precautions rooms only) | 108 sq in |
| Door touch areas | High touch areas of door – handles, door grab areas (relevant precautions rooms only) | 130 sq in |
| Sinks | Sink area – sink top, handles, soap dispenser button, hand sanitizer dispenser button | 87 sq in |
| Shared unit equipment | Surfaces of reusable shared unit equipment used in patient rooms | Variable |
| Shared nursing area | Composite of high touch areas in nurses station – two computer keyboards and two telephone handles | 140 sq in |

### Section 2: Microbiologic methods

To specifically isolate presumptive vancomycin-resistant *Enterococcus* strains, methods for sponge samples and swab samples vary slightly, but both involved culturing on VRE Chrome ID agar plates or in Bile Esculin Broth (BEA) with a vancomycin disk. Sponges were processed in a stomacher bag with 45 mL 0.02% Tween 80 buffer and the sponge eluate concentrated by centrifugation. To quantify VRE, 200 µL of each sponge eluate was pipetted onto VRE Chrome ID agar plates. For broth enrichment of the eluate, 500 µL was dispensed into bile esculin azide (BEA) broth with a 30 µg vancomycin disk. For swab samples, 100 µL of the vortexed swab transport media was plated onto VRE Chrom ID plates and 100 µL dispensed into BEA broth with a 30 µg vancomycin disk.

Plates were checked for growth at 24 and 48 hours, and the corresponding broth checked at 48 hours if no enterococci growth was seen on a plate. Plates and broths were held for 72 hours before discarding. If enterococci growth was observed on VRE Chrome ID plates, colony counts were recorded after 48 hours, and morphologically-distinct suspected enterococci colonies were subcultured to blood agar for identification by MALDI-TOF (Bruker Biotyper CA System, Bellerica, MA). If VRE Chrome ID agar plates were negative for enterococci at 48 hours, but growth was observed in the bile esculin broth with vancomycin disk, broth was streaked onto VRE Chrome ID agar plates for confirmation.

### Section 3: Alignment-based genomic methods

Raw reads were trimmed for adapters and contaminants using bbduk from bbmap (version 39.10)^37^ and then quality-checked using fastqc (version v0.12.1).^38^ We used SPAdes (version v4.2.0)^39^ to assemble contig short reads in full and plasmid assemblies. To identify specific resistance and virulence genes (Table S-3), full and plasmid assemblies were annotated using bakta (version 1.12.0)^40^ against the full database version 6.0.^41^ To identify the clade of each VRE isolate, assemblies were run against reference genomes for each clade A1, A2, and B (Table S-3) using fastANI (version 1.34)^25^ to determine the closest match. We used snippy (version 4.6.0)^42^ to create a single core alignment of each isolate against their respective reference genomes. To account for recombination, we ran gubbins (version 3.4)^43^ to establish a phylogeny of all isolates and then ran snp-dists (version 0.8.2)^44^ to establish pairwise single nucleotide variants (SNVs).

### Section 4: Genomic clustering sensitivity analysis

All methods were used with both SNV thresholds commonly used in the literature: 7^12^ and 20.^9,34^ All combinations of methods, parameters, and SNV thresholds were included in the sensitivity analysis (Table S-5). Any combination of methods, parameters, and SNV thresholds that produced different clusters was reported.

**Table S-3.** Key *Enterococcus* resistance and virulence factor genes.

| **Category** | **Gene** | **Function** | **Source** |
|---|---|---|---|
| Resistance | *vanA, (vanS-A, vanR-A, vanY-A, vanX-A, vanH-A)* | Vancomycin resistance | <sup>45</sup> |
| Resistance | *vanB* | Vancomycin resistance | <sup>45</sup> |
| Resistance | *vanZ* | Teicoplanin resistance | <sup>45</sup> |
| Virulence factor | *ace*/*acm* | Collagen adhesion | <sup>46</sup> |
| Virulence factor | *asa1* | Adhesion | <sup>46</sup> |
| Virulence factor | *cylA* | Cytolysin toxin | <sup>46</sup> |
| Virulence factor | *efaA* | Cell surface antigen | <sup>46</sup> |
| Virulence factor | *esp* | Enterococcal surface protein, biofilm formation | <sup>46,47</sup> |
| Virulence factor | *gelE* | Gelatinase, biofilm formation | <sup>46</sup> |
| Virulence factor | *hyl* | Hyaluronidase, tissue invasion | <sup>46</sup> |

**Table S-4.** Reference genomes for each VRE *Enterococcus faecium* clade.

| **Clade** | **Reference name** | **NCBI RefSeq assembly** |
|---|---|---|
| A1 | AUS0004 | GCF_000250945.2 |
| A2 | F88 | GCF_019175425.1 |
| B | BIOPOP3-WT | GCF_012045505.1 |

**Table S-5.** List of competing methods and parameters for the genomic clustering sensitivity analysis.

| Genomic Method | Parameters | Reference for parameters |
|---|---|---|
| Split-kmer analysis (SKA) | default | <sup>12</sup> |
| Split-kmer analysis (SKA) | optimized | <sup>28</sup> |
| Alignment-based methods | Not applicable | Not applicable |

### Section 5: Results

#### Sample Collection

While the total number of samples collected from the four main sampling sources (patients, HCP hands, patient room environments, and shared environments) totaled 6848, there were additional samples that were collected for quality control. Overall, the total number of samples tested for microbiologic detection was 7000. These included controls and samples from the sampler hands. These went through identical microbiologic testing to ensure quality control and rule out laboratory contamination. None of the controls or sampler hand samples yielded VRE colonies, so they are not included in our analysis.

#### Genomic analyses

The *Enterococcus faecium* core genome alignment generated by snippy against the AUS0004 reference genome comprised 3,027,337 bp. After removal of recombinant regions with gubbins, 925 polymorphic sites remained and were used for phylogenetic reconstruction and pairwise SNV distance calculations.

**Figure S-1.**
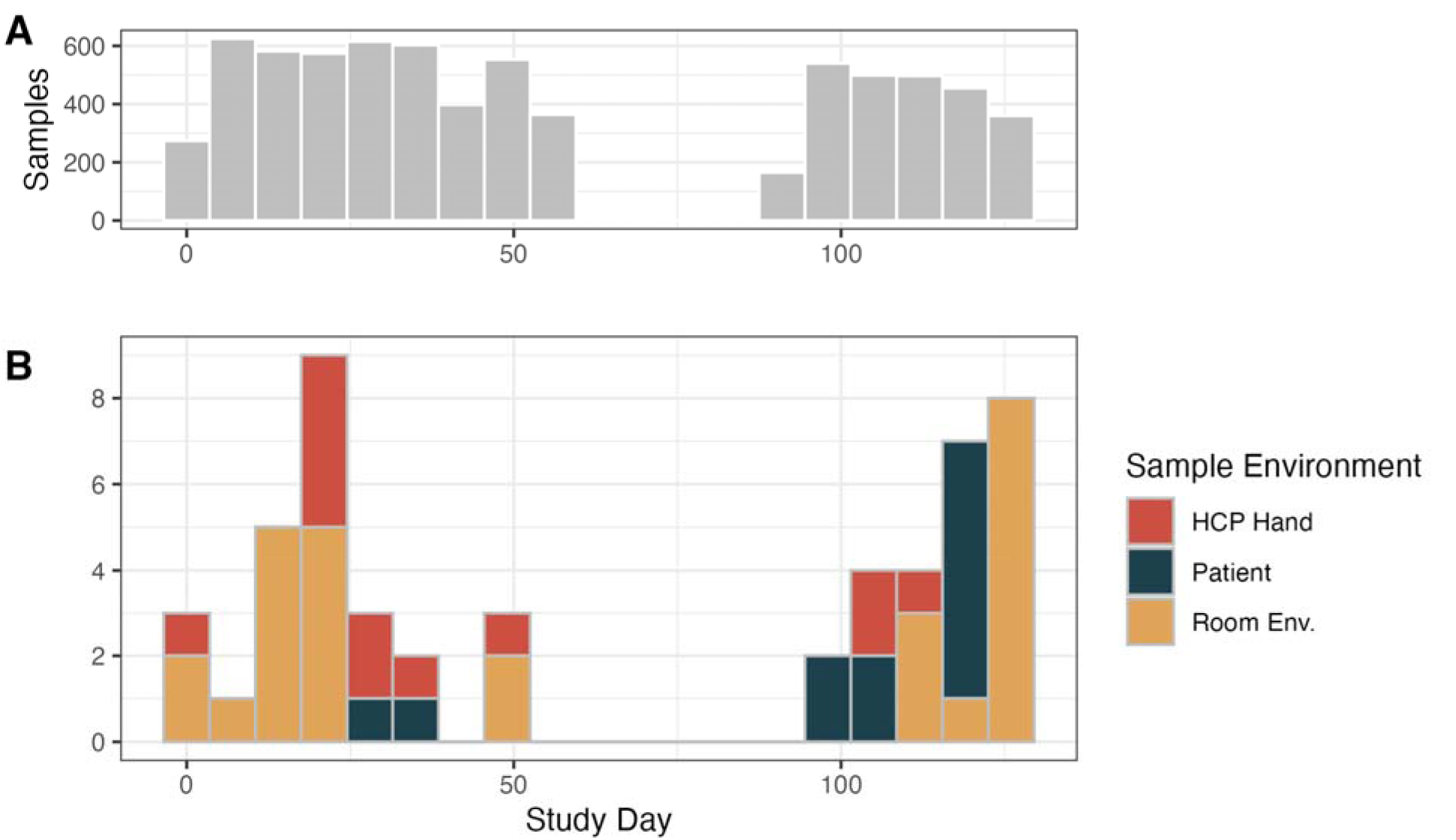
Study timeline of sampling effort and vancomycin-resistant Enterococcus (VRE) detection. A) Histogram of the total number of samples collected each week, with hospital A samples collected days 1-56 and hospital B samples collected days 93-127. B) Histogram of all samples yielding one or more VRE isolate over time, colored by the sample environment. Since no VRE was isolated from the shared environmental surfaces, it is omitted from this histogram.

**Table S-6.**
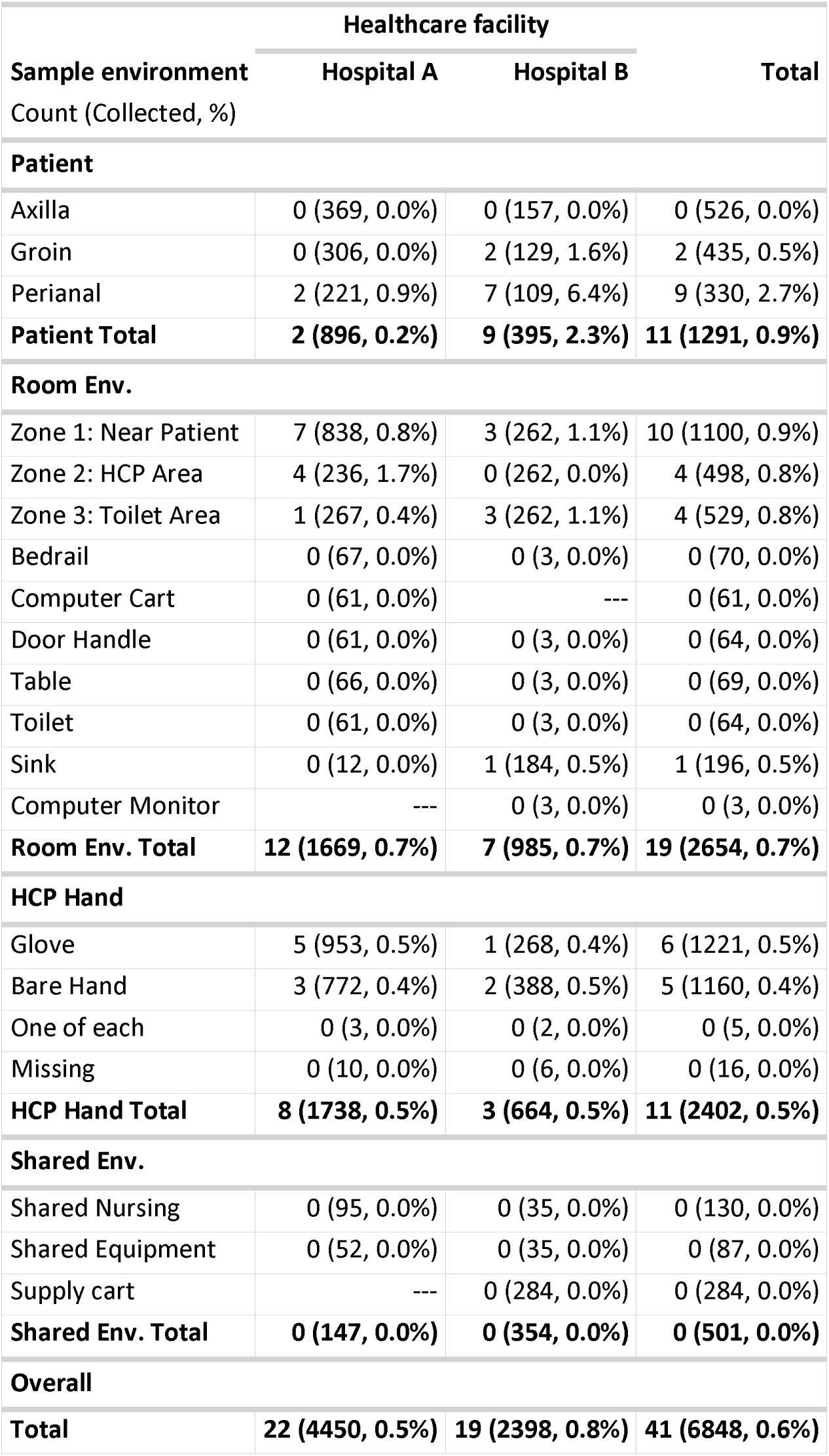
Count of samples yielding at least one VRE isolates (Number of samples collected, %), by facility and sampling environment.

|  | **Healthcare facility** |  |  |
|---|---|---|---|
| **Sample environment**<br>Count (Collected, %) | **Hospital A** | **Hospital B** | **Total** |
| **Patient** |  |  |  |
| Axilla | 0 (369, 0.0%) | 0 (157, 0.0%) | 0 (526, 0.0%) |
| Groin | 0 (306, 0.0%) | 2 (129, 1.6%) | 2 (435, 0.5%) |
| Perianal | 2 (221, 0.9%) | 7 (109, 6.4%) | 9 (330, 2.7%) |
| **Patient Total** | **2 (896, 0.2%)** | **9 (395, 2.3%)** | **11 (1291, 0.9%)** |
| **Room Env.** |  |  |  |
| Zone 1: Near Patient | 7 (838, 0.8%) | 3 (262, 1.1%) | 10 (1100, 0.9%) |
| Zone 2: HCP Area | 4 (236, 1.7%) | 0 (262, 0.0%) | 4 (498, 0.8%) |
| Zone 3: Toilet Area | 1 (267, 0.4%) | 3 (262, 1.1%) | 4 (529, 0.8%) |
| Bedrail | 0 (67, 0.0%) | 0 (3, 0.0%) | 0 (70, 0.0%) |
| Computer Cart | 0 (61, 0.0%) | --- | 0 (61, 0.0%) |
| Door Handle | 0 (61, 0.0%) | 0 (3, 0.0%) | 0 (64, 0.0%) |
| Table | 0 (66, 0.0%) | 0 (3, 0.0%) | 0 (69, 0.0%) |
| Toilet | 0 (61, 0.0%) | 0 (3, 0.0%) | 0 (64, 0.0%) |
| Sink | 0 (12, 0.0%) | 1 (184, 0.5%) | 1 (196, 0.5%) |
| Computer Monitor | --- | 0 (3, 0.0%) | 0 (3, 0.0%) |
| **Room Env. Total** | **12 (1669, 0.7%)** | **7 (985, 0.7%)** | **19 (2654, 0.7%)** |
| **HCP Hand** |  |  |  |
| Glove | 5 (953, 0.5%) | 1 (268, 0.4%) | 6 (1221, 0.5%) |
| Bare Hand | 3 (772, 0.4%) | 2 (388, 0.5%) | 5 (1160, 0.4%) |
| One of each | 0 (3, 0.0%) | 0 (2, 0.0%) | 0 (5, 0.0%) |
| Missing | 0 (10, 0.0%) | 0 (6, 0.0%) | 0 (16, 0.0%) |
| **HCP Hand Total** | **8 (1738, 0.5%)** | **3 (664, 0.5%)** | **11 (2402, 0.5%)** |
| **Shared Env.** |  |  |  |
| Shared Nursing | 0 (95, 0.0%) | 0 (35, 0.0%) | 0 (130, 0.0%) |
| Shared Equipment | 0 (52, 0.0%) | 0 (35, 0.0%) | 0 (87, 0.0%) |
| Supply cart | --- | 0 (284, 0.0%) | 0 (284, 0.0%) |
| **Shared Env. Total** | **0 (147, 0.0%)** | **0 (354, 0.0%)** | **0 (501, 0.0%)** |
| **Overall** |  |  |  |
| **Total** | **22 (4450, 0.5%)** | **19 (2398, 0.8%)** | **41 (6848, 0.6%)** |

**Figure S-2.**
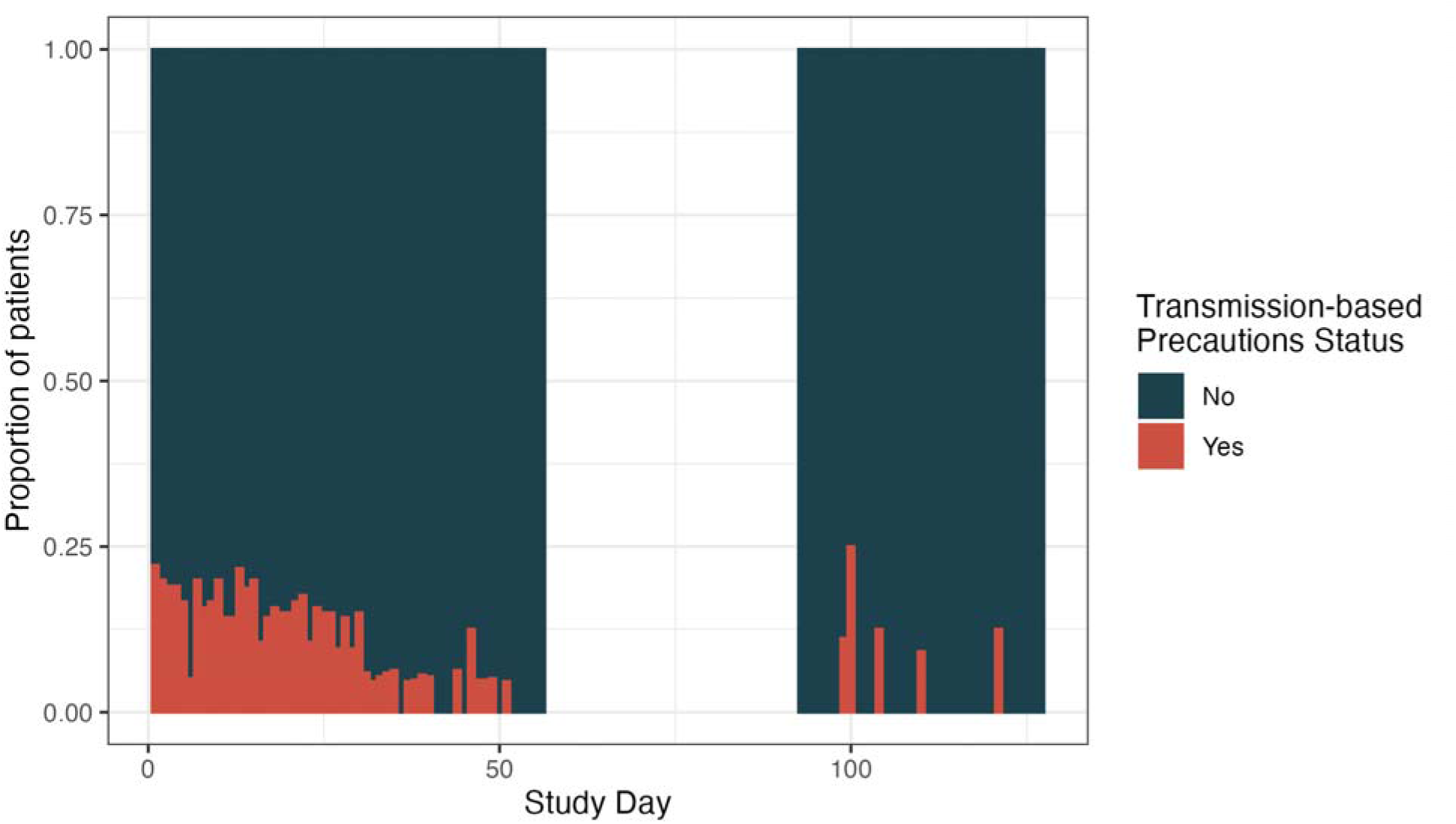
Proportion of patients in hospital A’s CV-ICU (study days 1-56) and hospital B’s ICU (study days 93-127) on any type of transmission-based precautions.

**Figure S-3.**
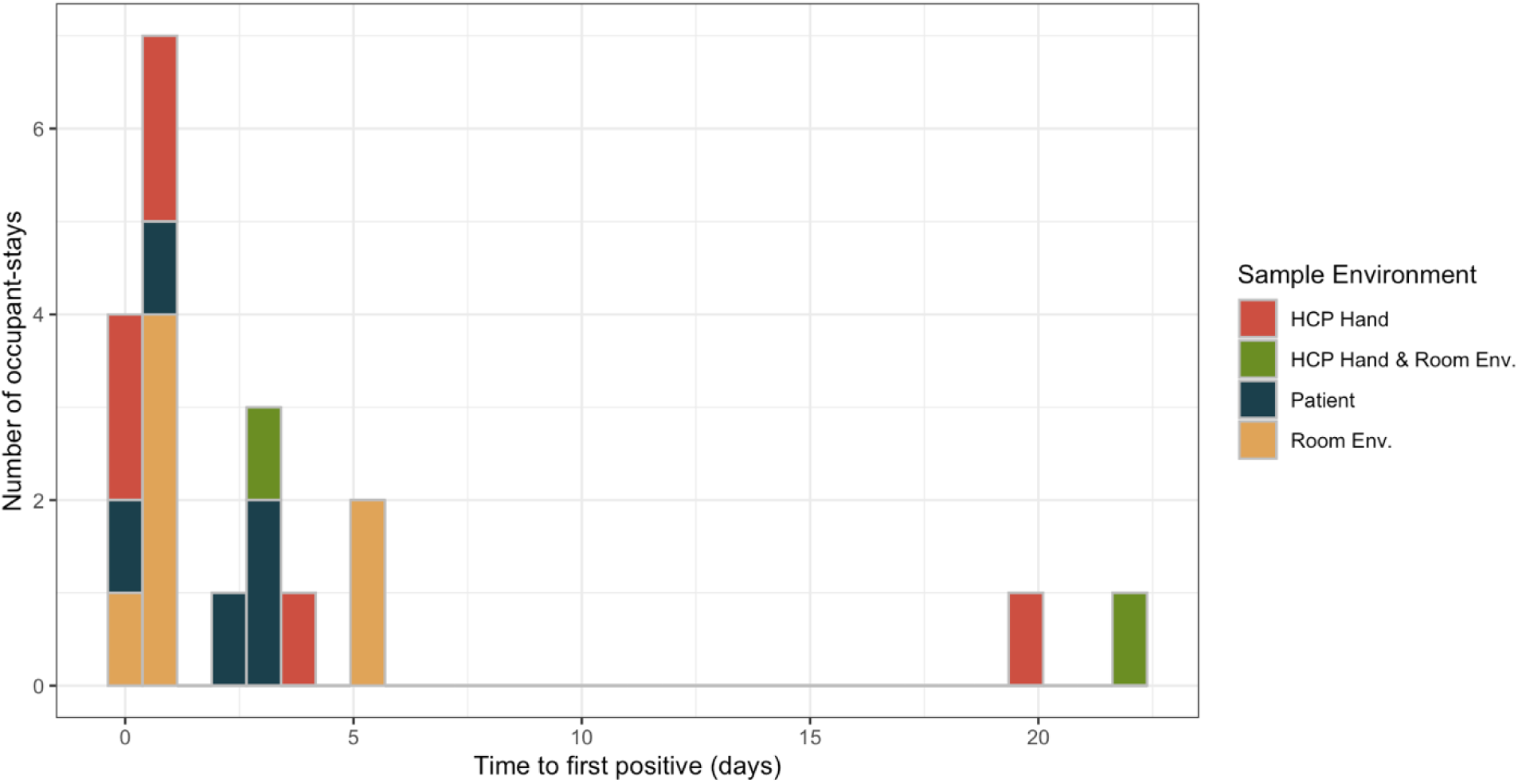
Histogram of occupant-stays with VRE detection with the time (in days) between patient admission and VRE detection, with fill color representing the sample environment that first yielded VRE.

**Figure S-4.**
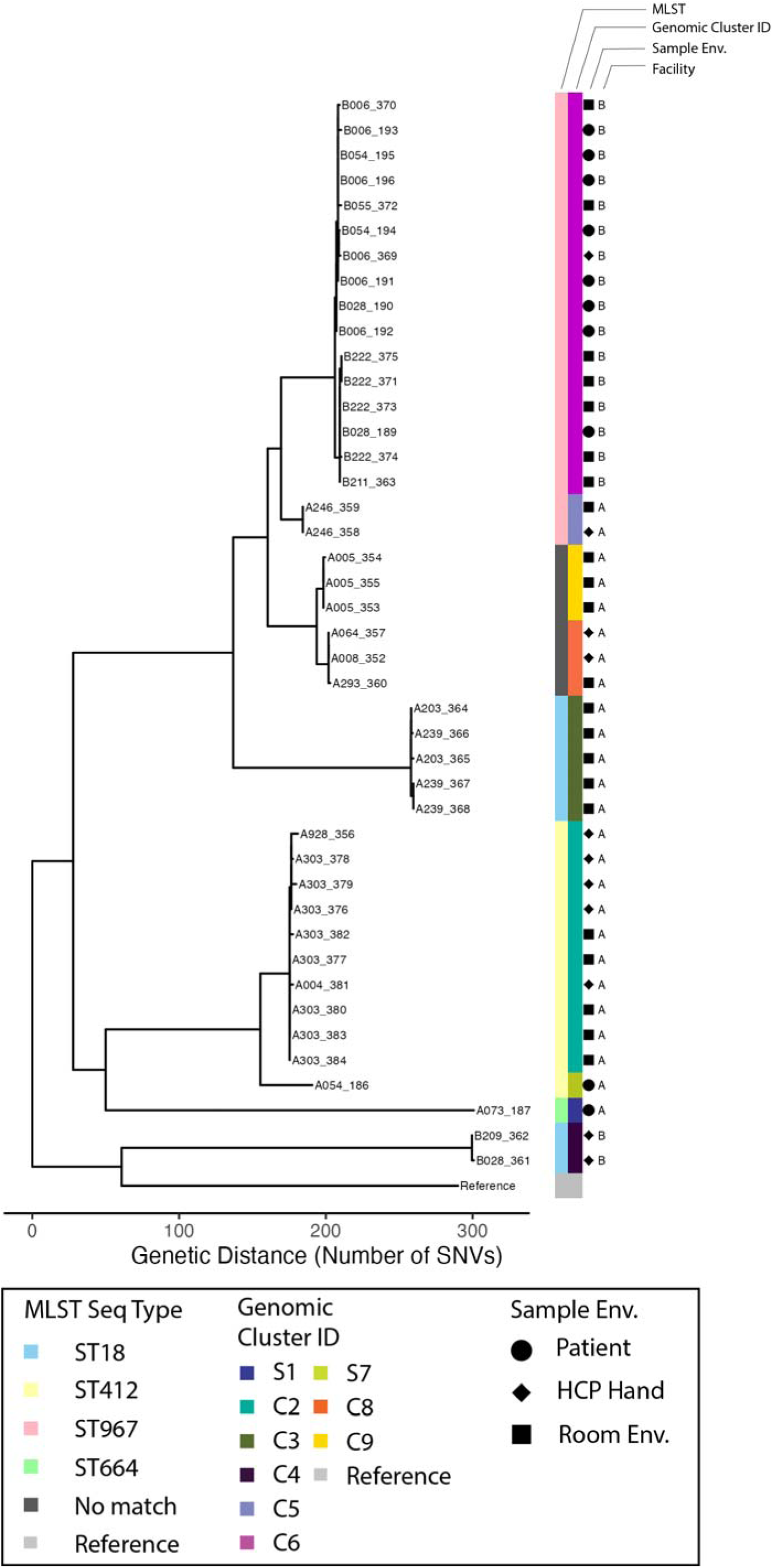
Maximum likelihood phylogenetic tree of all vancomycin-resistant Enterococcus faecium (VREfm) isolates generated after alignment against the Clade A1 reference genome (AUS004) and recombinant region filtering. Tips are labeled with the occupant-stay identifier that their samples are affiliated with, followed by the whole genome sequence identifier (e.g., A303_384). Metadata associated with each tip include the multilocus sequence types (MLST) of each isolate when available, the unique Cluster ID as identified via our clustering algorithm, the sample environment, and the healthcare facility.

**Figure S-5.**
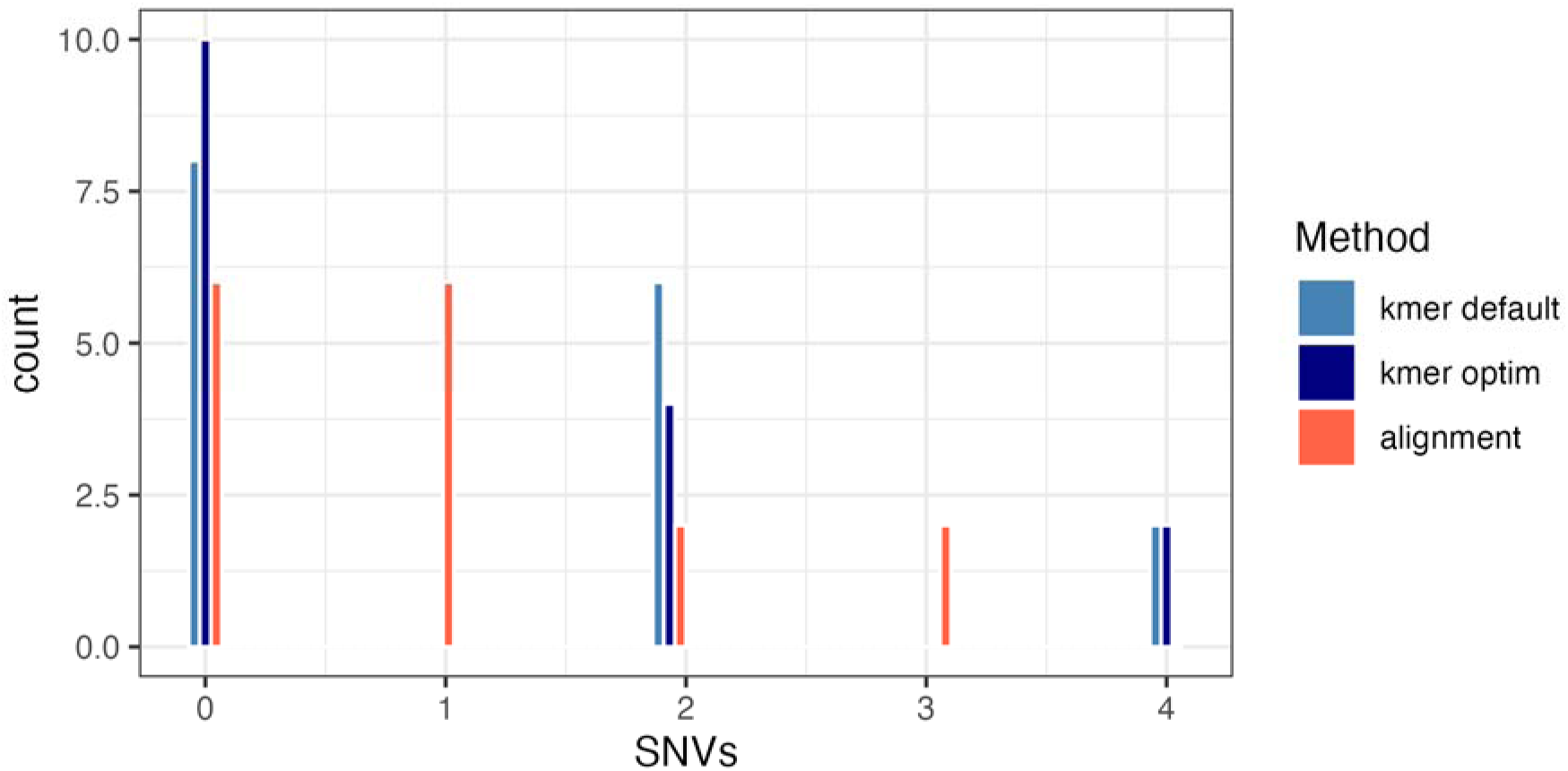
Histogram of the pairwise SNV-distances calculated between all pairs of VRE isolates taken from the same patient, body site samples only. The pairwise SNVs calculated using each method are depicted by the color of the bar, with kmer-based methods using different parameters in blue, and the alignment based method in red.

**Figure S-6.**
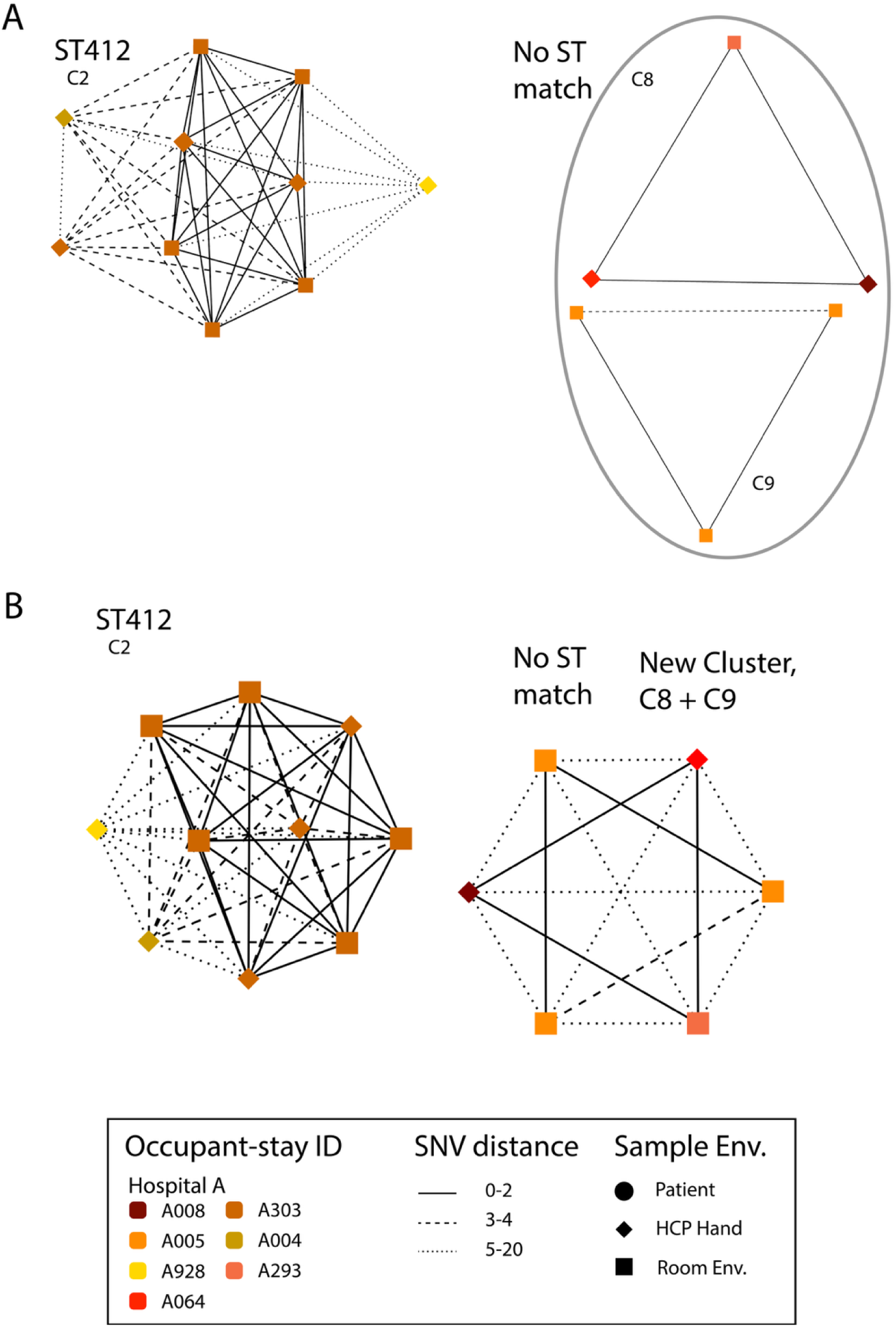
Network plots of VREfm (vancomycin-resistant Enterococcus faecium) multi-isolate clusters from the single nucleotide variant (SNV) threshold sensitivity analysis. A) Network plots of VREfm clusters using the 7 SNV pairwise distance threshold for clustering links. B) Network plots of VREfm clusters using a 20 SNV pairwise distance threshold for clustering links. Nodes represent individual VREfm isolates, which are colored based on the occupant-stay that the sample was affiliated with, and with shapes based on the sample environment. Edges that connect each node are weighted by the pairwise SNV distances. Only clusters that changed from the 7 to the 20 SNV thresholds are depicted. Clusters are labeled by their matching multi-locus sequence type. Sequence types with more than one cluster are circled in gray.

